# Hyperlipidemia Pharmacotherapy in Skilled Nursing Facilities: A Real-World Evidence Study

**DOI:** 10.64898/2026.06.11.26355474

**Authors:** Huda Ashraf, Katherine E. Mathers, Brittin Wagner, Tyler M. Saumur

## Abstract

**Objectives:** To estimate hyperlipidemia medication order prevalence and associated variables in U.S. skilled nursing facility (SNF) residents.

**Design:** Retrospective, observational study.

**Setting and Participants:** Electronic Health Record data from 447,080 SNF residents with a hyperlipidemia diagnosis identified in PointClickCare’s Life Sciences clinical database (January-April 2025) were reviewed.

**Methods:** The presence and absence of medication orders for hyperlipidemia treatments recommended by the American Heart Association were assessed. Descriptive analyses summarized demographic and clinical characteristics, and a modified Poisson regression model was used to estimate risk ratios for having a medication order, adjusting for demographic, clinical, and facility characteristics.

**Results:** Overall, 83.3% of residents diagnosed with hyperlipidemia had at least one hyperlipidemia medication order. Statins were ordered by 96.2% of active order residents, while other medication classes i.e., omega-3 fatty acids, cholesterol absorption inhibitors, fibrates were less common (<8%). Risk ratios (RRs) for medication orders ranged from 0.87-1.16. Factors most strongly associated with having an order included hypertension medication orders (RR=1.16), unspecified hyperlipidemia diagnosis (RR=1.10), and active diabetes medication orders (RR=1.09); female sex (RR=0.95) and private (0.94) or other (0.87) payer types were associated with a lower likelihood of having an order.

**Conclusions and Implications:** Most residents with a hyperlipidemia diagnosis had an active relevant medication order, but use of non-statin therapies was rare. Differences in treatment patterns by sex and payer type, along with limited uptake of newer agents, warrant further investigation into prescribing practices and access within SNFs.

**Brief Summary:** Most SNF residents with hyperlipidemia had relevant medication orders; non-statin use was rare, and differences observed in prescribing patterns by sex and payer warrant further study.

## Introduction

Hyperlipidemia is a prevalent condition in the United States (U.S.), affecting nearly 95 million adults and contributing significantly to the burden of atherosclerotic cardiovascular disease (ASCVD), the leading cause of death among older adults.^1,2^ Although statin therapy has been shown to reduce major cardiovascular events and mortality even among those aged 65 years and older, treatment rates remain inadequate. Among U.S. adults who meet criteria for statin therapy, less than half are actively taking the medication.^3,4,5^ In high-risk clinical populations such as those with comorbid coronary artery disease and heart failure, lipid-lowering medication use has been shown to be lower than those with coronary artery disease alone.^6^ Similar patterns of low treatment rates have also been observed in the long-term care (LTC) setting and those who have been discharged following acute coronary events.^7, 8^ These patterns in treatment appear to be driven in part by concerns related to advanced age, comorbidity burden, polypharmacy, and drug-drug interactions, particularly in older and LTC populations.^9, 10^

Recent U.S. data reveal persistent treatment gaps in lipid management regarding specific demographic characteristics. Among adults with atherosclerotic cardiovascular disease, women are significantly less likely than men to receive statin therapy.^11^ Markedly lower statin use has also been observed among racial and ethnic minorities at risk of developing ASCVD; with treatment rates of 20% in Black adults and 15.4% in Hispanic adults compared with 27.9% in White adults.^12^ National data further show that non-Hispanic Black men and Mexican-American women remain significantly undertreated compared with non-Hispanic White men.^13^ For secondary prevention, the use of guideline-recommended lipid-lowering therapies also remains suboptimal, indicating persistent barriers to the implementation of evidence-based recommendations in clinical practice.^14^ Such demographic disparities highlight missed opportunities to reduce preventable cardiovascular events across U.S. populations.

While prior studies have reported treatment gaps, largely in community-dwelling populations, there is limited, contemporary, large-scale evidence describing treatment patterns in skilled nursing facilities and the drivers of treatment. This study addresses these gaps by leveraging a large, national EHR dataset to describe contemporary lipid-lowering medication order patterns among U.S. SNF residents with hyperlipidemia and to identify resident- and facility-level factors associated with having an active order.

## Methods

### Setting and Participants

This retrospective, cross-sectional observational study utilized deidentified electronic health record (EHR) data from the PointClickCare Life Sciences database, one of the largest real-world datasets of U.S. LTC residents. The database includes resident demographics, Minimum Data Set clinical and functional assessments, medication and vaccination records, vital signs, and related data elements from over 18 million residents. Data is deidentified and expert-determined from residents with facility-obtained consent and covered by business associate agreements with LTC facilities. Exemption from ethics committee oversight was granted by the Research Ethics Boards of the University of Toronto and University of Alberta.

During the study period (January–April 2025), EHR data were available for 1,285,062 individuals within assisted living, independent living, and skilled nursing facilities (SNFs). Among these, 447,080 residents in SNFs had a documented diagnosis of hyperlipidemia, identified using ICD-10 codes E78.2 (mixed hyperlipidemia) and E78.5 (unspecified hyperlipidemia). Residents were included if they had at least one hyperlipidemia diagnosis and at least one day of SNF residence during the study window and were active in the facility at the study end date. Hyperlipidemia subtype categories were not mutually exclusive; some residents had more than one diagnosis code during the study window.

### Exposures and Variables of Interest

Residents were categorized based on the presence or absence of medication orders for hyperlipidemia treatments consistent with American Heart Association guidelines.^15^ Treatment included statins, cholesterol absorption inhibitors, omega-3 fatty acids, fibrates, bile acid sequestrants, nicotinic acid derivatives, and PCSK9 inhibitors.

Resident-level variables included demographic characteristics (age, sex, race/ethnicity), insurance status, facility region, and length of stay. Days in facility (DIF) was calculated as the number of days between a resident’s admission and discharge dates (for residents in facility at the end of the study period, 30 April 2025 was used as the discharge date. Consecutive visits were combined when separated by gaps of seven days or fewer, treating them as a single continuous stay for DIF calculation). Diagnoses for clinical comorbidities such as diabetes and hypertension were included due to their potential influence on medication order patterns.

### Data Analysis

Data were extracted using SQL and analyzed in Python. Descriptive statistics were used to summarize demographics and treatment characteristics. A modified Poisson regression model was fit with a log link to estimate prevalence risk ratios (RRs) and 95% confidence intervals (CIs). Covariates included resident demographics, clinical characteristics, functional and cognitive measures, facility characteristics, payer type, and area-level socioeconomic indicators. Categorical variables were dummy-coded with pre-specified reference categories: female sex, White race, Medicare fee-for-service payer, South region, and non-profit facility ownership. Continuous variables were rescaled prior to model entry to improve interpretability of effect estimates: age was expressed per 10-year increment, BMI per 5-unit increment, number of facility beds per 25-bed increment, and DIF per 30-day increment. Statistical significance was assessed at α = 0.001.

## Results

There were 447,080 residents with hyperlipidemia who met the eligibility criteria (Table 1). Within this cohort, 83.3% had at least one active hyperlipidemia medication order. Across this population, the mean (SD) age was 77.0 (11.7) years, with the majority of residents being female (58.6%) and White (67.6%). Residents were distributed across

**Table 1.**
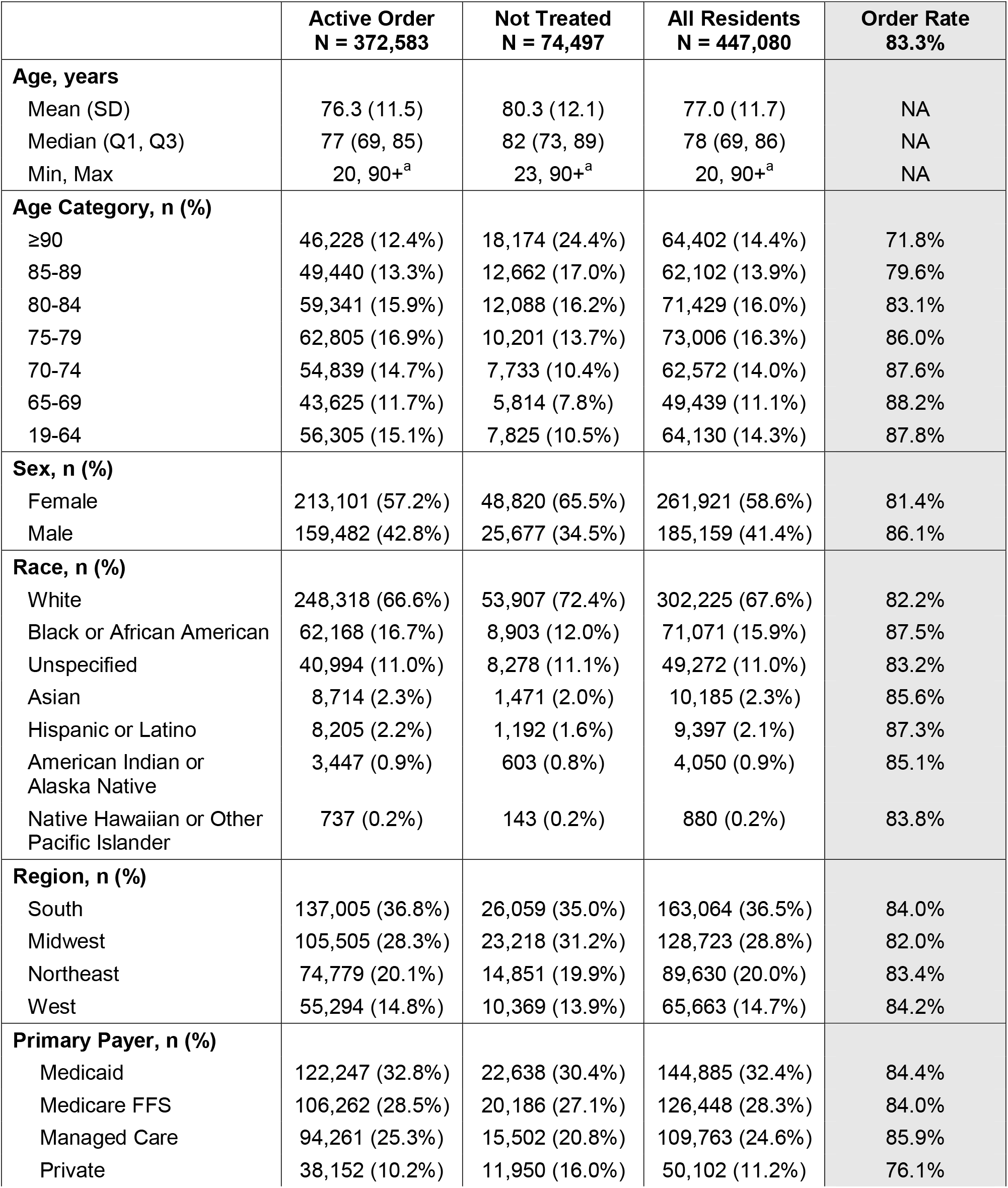
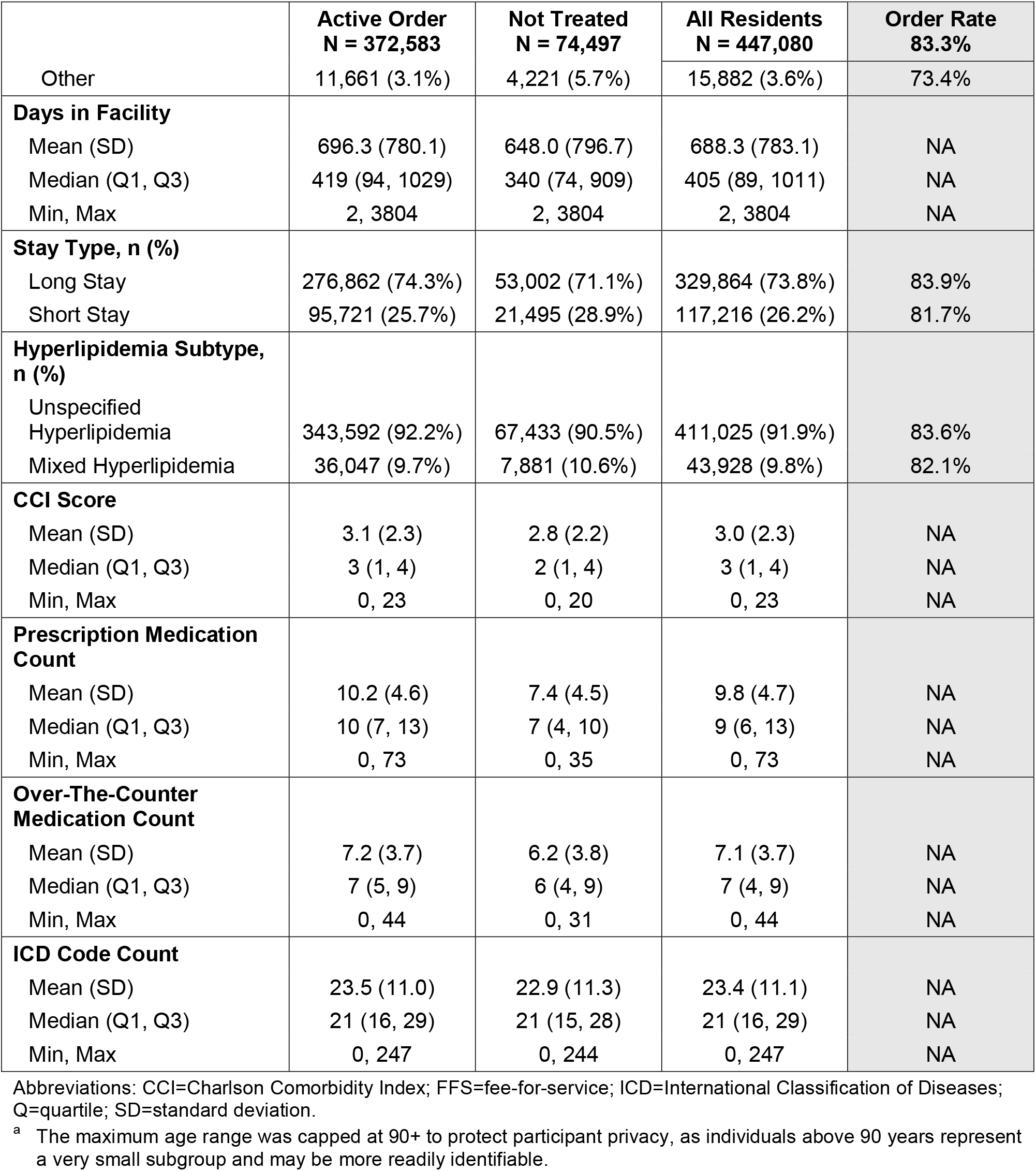
Demographic and Clinical Characteristics Across Exposure Groups.

U.S. regions, with the South (36.5%) and Midwest (28.8%) most represented. Primary payer types included Medicaid (32.4%), Medicare fee-for-service (28.3%), and Managed Care (24.6%), with smaller proportions covered by private insurance and other sources.

The majority of residents had unspecified hyperlipidemia (91.9%), with mixed hyperlipidemia present in 9.8%. Long-stay residents comprised 73.8% of the cohort. Among active order residents, statins were the most commonly ordered medication class (96.2%), followed by omega-3 fatty acids (7.0%), with all other medications under 5.0% (Table 2).

**Table 2.** Active Order Rates by Medication Class.

|  | Active Order<br>N = 372,583 |
| --- | --- |
| **Medication Class, n (%)** |  |
| Statin | 358,565 (96.2%) |
| Omega-3 Fatty Acid | 26,215 (7.0%) |
| Cholesterol Absorption Inhibitors | 16,315 (4.4%) |
| Fibrate | 16,139 (4.3%) |
| Bile Acid Sequestrant | 9,981 (2.7%) |
| Nicotinic Acid | 8,194 (2.2%) |
| PCSK9 Inhibitor | 1,000 (0.3%) |

Abbreviations: PCSK9=proprotein convertase subtilisin/Kexin type 9.

RRs for having hyperlipidemia medication orders were estimated using a modified Poisson regression model (Figure 1) and ranged from 0.87 to 1.16 (Supplementary Table 1). The strongest positive associations with medication orders were observed for hypertension medication use (RR = 1.16; 95% CI [1.15-1.16]), unspecified hyperlipidemia diagnosis (RR = 1.10 [1.10-1.11]), diabetes medication use (RR = 1.09 [1.09-1.10]), and mixed hyperlipidemia (RR = 1.08 [1.07-1.08]). Male sex (RR = 1.05 [1.05-1.05]) and Black race (RR = 1.04 [1.04-1.04]) were demographic features also associated with a higher likelihood of medication orders. In contrast, lower likelihoods of medication orders were observed for other payer type (RR = 0.87 [CI 0.86-0.88]), private payer status (RR = 0.94 [0.94-0.95]), age (RR = 0.97 [0.97-0.97]), Medicaid coverage (RR = 0.97 [0.97-0.98]), and Midwestern U.S. (RR = 0.97 [0.97-0.98]).

**Figure 1.**
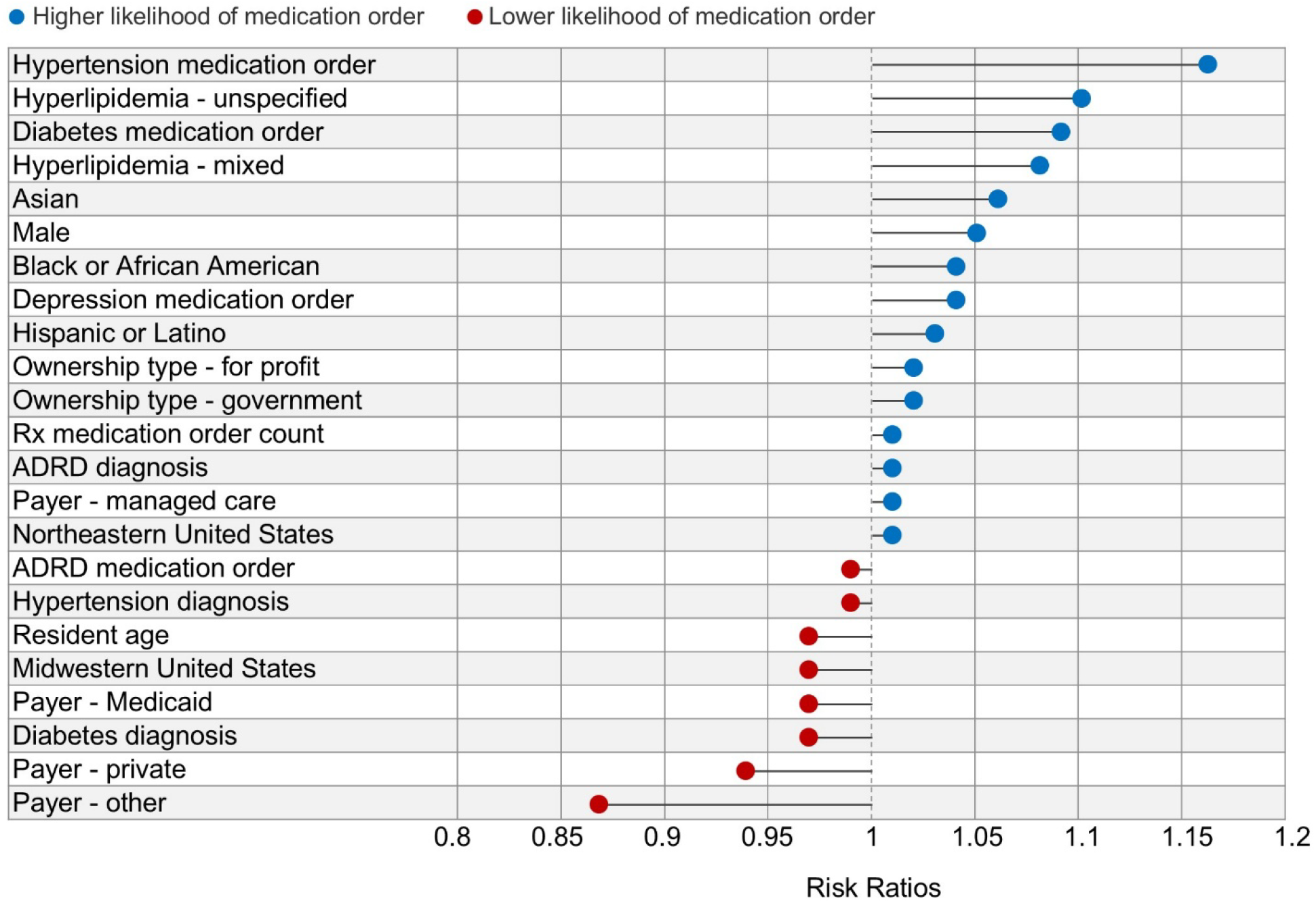
Summary of risk ratios. Abbreviations: ADRD=Alzheimer’s disease and related dementias; Rx=prescription. Note: The figure presents all variables in the model with risk ratios greater than or less than 1.00.

## Discussion

This study described lipid-lowering medication order patterns among a large national cohort of U.S. SNF residents with hyperlipidemia. Overall, 83.3% of residents diagnosed with hyperlipidemia had at least one active lipid-lowering medication order during the study window as recommended by the American Heart Association, with statins as the most commonly prescribed medications (96.2%). Use of non-statin agents such as cholesterol absorption inhibitors and fibrates was very low (~4% each), omega-3 fatty acids were used in 7.0% of residents, and PCSK9 inhibitors accounted for only 0.3% of prescriptions, despite their role in achieving lipid goals for high-risk patients. Residents taking diabetes medications had a higher likelihood of receiving lipid-lowering therapy, whereas female residents and those with private or other payer types were less likely to receive guideline-concordant care.^11,13^

The proportion of SNF residents receiving hyperlipidemia medication in this study is significantly higher than rates reported in community-dwelling older adults, where statin use remains suboptimal at <50% of those recommended for therapy based on cardiovascular risk or hyperlipidemia.^5^ While advances in lipid management and the introduction of newer agents have expanded therapeutic alternatives to statins, our findings suggest that variation in prescribing remains evident within SNF settings.

Factors unique to SNF, such as polypharmacy, clinical complexity, and competing clinical priorities, may contribute to undertreatment or therapeutic inertia.^10^ Interestingly, active order residents were prescribed more medications on average (10.2 vs 7.4 for untreated), indicating that a higher medication burden may actually increase the likelihood of receiving hyperlipidemia therapy. Notably, while taking diabetes medications was strongly associated with an active hyperlipidemia medication order, having a diabetes diagnosis was associated with a lower likelihood, highlighting a disconnect between diagnosis and management.

The low uptake of non-statin therapies, despite guideline recommendations, highlights ongoing challenges in translating evidence into practice for vulnerable populations.^14^ These findings align with national trends showing that, despite increased statin use over time, expenditures and adoption of newer lipid-lowering agents remain limited, particularly among older adults.^16^ The low prevalence of non-statin therapy orders may reflect a combination of prescribing preferences, formulary access, cost barriers, or limited perceived benefit in this population.

Differences in prescribing patterns were observed by sex, payer, and region. These differences may reflect variation in resident case mix, prescribing practices, drug coverage, or facility-level access rather than a single underlying mechanism. Female residents were modestly less likely to have an active hyperlipidemia compared to male residents, echoing prior research documenting lower prescription rates among women.^11^ Regional and socioeconomic differences, with lower likelihoods of treatment in the Midwest and among residents with private or other payer types, may reflect variations in prescriber practice, formulary access, resource availability, or systemic barriers at the facility level. These findings underscore the need to further explore individual- and system-level determinants of care.^13^

Our results also revealed that residents with diabetes medications and other chronic disease medications were more likely to receive lipid-lowering therapy, whereas older residents and those with certain payer types were less likely to have active medication orders. This pattern may reflect greater engagement with healthcare among residents on chronic medications, or alternatively, competing clinical priorities and concerns about adverse effects in more complex patients.^11^ The heavy reliance on statins, coupled with minimal use of adjunctive therapies, suggests that prescriber familiarity and formulary constraints may influence treatment decisions more than patient-specific lipid goals.

Several limitations should be considered. First, the cross-sectional design precludes causal inference, and unmeasured confounding variables may have influenced the observed associations. In addition, medication history before the study window was not considered and those who discontinued hyperlipidemia treatment are not captured as ‘treated’ in the present study. Second, the analysis was limited to pharmacologic medication orders and did not capture non-medication interventions such as dietary management or physical activity. The capture of treatment rates through medication orders may also artificially increase the proportion receiving treatment as administration information was not captured. Third, findings may not be generalizable beyond SNF settings or to healthcare systems with different prescribing practices. Nevertheless, this study leverages a large, real-world EHR dataset and provides important insights into current patterns of hyperlipidemia care in SNF.^3^

## Conclusions and Implications

The findings of this study reveal potential gaps in the delivery of pharmacological hyperlipidemia treatment among SNF residents, with differences affecting female residents and various payer types. Although active hyperlipidemia medication orders were common, the limited use of non-statin, evidence-based therapies such as cholesterol absorption inhibitors, omega-3 fatty acids, and PCSK9 inhibitors indicates that access to recommended medications remains uneven across clinical and demographic groups. The increased likelihood an active order among residents with diabetes and those already receiving chronic disease medications suggests that greater medical oversight and comorbidity management may improve identification and management of hyperlipidemia. Future work should identify strategies to improve prescribing and access to advanced therapies in LTC settings.

## Data Availability

Data produced in the present study are available upon reasonable request to the authors.

## Acknowledgements

We would like to thank Jody Long and Dr. Steve Buslovich for their collaboration and their clinical input provided for this manuscript. We would also like to thank Michelle Sweeny, Aaron Norfolk, and Kaitlyn Riddell for providing their perspectives. This research was supported by PointClickCare Life Sciences and McMaster University. McMaster University Library provided journal article access.

## References

1. Grundy SM, Stone NJ, Bailey AL, et al. 2018 AHA/ACC/AACVPR/AAPA/ABC/ACPM/ADA/AGS/APhA/ASPC/NLA/PCNA guideline on the management of blood cholesterol: a report of the American College of Cardiology/American Heart Association Task Force on Clinical Practice Guidelines. Circulation. 2019;139(25):e1082–e1143.

2. Szadkowska I, Stanczyk A, Aronow WS, et al. Statin therapy in the elderly: a review. Arch Gerontol Geriatr. 2010;50(1):114–8.

3. Alexander GC, Curran J, Victores A, et al. US public health gains from improved treatment of hypercholesterolemia: a simulation study of NHANES adults treated to guideline-directed therapy. J Gen Intern Med. 2025. [add pages/DOI when available]

4. Sayed A, Navar AM, Slipczuk L, et al. Prevalence, awareness, and treatment of elevated LDL cholesterol in US adults, 1999–2020. JAMA Cardiol. 2023;8(12):1185–94.

5. Thompson-Paul AM, Gillespie C, Wall HK, et al. Recommended and observed statin use among U.S. adults – National Health and Nutrition Examination Survey, 2011–2018. J Clin Lipidol. 2023;17(2):225–35.

6. Sueta CA, Massing MW, Chowdhury M, et al. Undertreatment of hyperlipidemia in patients with coronary artery disease and heart failure. J Card Fail. 2003;9(1):36–41.

7. Mack DS, Hume AL, Tjia J, Lapane KL. National trends in statin use among the United States nursing home population (2011–2016). Drugs Aging. 2021;38(5):427– 39.

8. Leya M, Stone NJ. Statin prescribing in the elderly: special considerations. Curr Atheroscler Rep. 2017;19(11):47.

9. Majumdar SR, Gurwitz JH, Soumerai SB. Undertreatment of hyperlipidemia in the secondary prevention of coronary artery disease. J Gen Intern Med. 1999;14(12):711–7.

10. Nair AP, Darrow B. Lipid management in the geriatric patient. Endocrinol Metab Clin North Am. 2009;38(1):185–206.

11. Shahid I, Satish P, Gullapelli R, et al. Gender disparities in utilization of statins for low density lipoprotein management across the spectrum of atherosclerotic cardiovascular disease: insights from the Houston Methodist Cardiovascular Disease Learning Health System Registry. Am J Prev Cardiol. 2024;19:100722.

12. Jacobs JA, Addo DK, Zheutlin AR, et al. Prevalence of statin use for primary prevention of atherosclerotic cardiovascular disease by race, ethnicity, and 10-year disease risk in the US. JAMA Cardiol. 2023;8(5):443–52.

13. Frank DA, Johnson AE, Hausmann LRM, Gellad WF, et al. Disparities in guideline-recommended statin use for prevention of atherosclerotic cardiovascular disease by race, ethnicity, and gender. Ann Intern Med. 2023;176(8):1057–66.

14. Navar AM, Kolkailah AA, Gupta A, et al. Gaps in guideline-based lipid-lowering therapy for secondary prevention in the United States: a retrospective cohort study of 322 153 patients. Circ Cardiovasc Qual Outcomes. 2023;16(8):533–43.

15. Cholesterol medications. American Heart Association. https://www.heart.org/en/health-topics/cholesterol/prevention-and-treatment-of-high-cholesterol-hyperlipidemia/cholesterol-medications. Accessed June 11, 2026.

16. Salami JA, Warraich H, Valero-Elizondo J, et al. National trends in statin use and expenditures in the US adult population from 2002 to 2013: insights from the Medical Expenditure Panel Survey. JAMA Cardiol. 2017;2(1):56–65.

