## Supplementary Table 1 for "Hyperlipidemia Pharmacotherapy in Skilled Nursing Facilities: A Real-World Evidence Study"

| **Supplementary Table 1 Complete List of Variables and Risk Ratios in the Modified Poisson Regression Model** | | |
| --- | --- | --- |
| **Variable** | **Risk Ratio [95% CI]** | **P-value** |
| Active hypertension medication order | 1.16 [1.15, 1.16] | <0.001 |
| Active hyperlipidemia - unspecified (E78.5) diagnosis | 1.10 [1.10, 1.11] | <0.001 |
| Active diabetes medication order | 1.09 [1.09, 1.10] | <0.001 |
| Active hyperlipidemia - mixed (E78.2) diagnosis | 1.08 [1.07, 1.08] | <0.001 |
| Asian | 1.06 [1.05, 1.07] | <0.001 |
| Male | 1.05 [1.05, 1.05] | <0.001 |
| Black or African American | 1.04 [1.04, 1.04] | <0.001 |
| Active depression medication order | 1.04 [1.03, 1.04] | <0.001 |
| Hispanic or Latino | 1.03 [1.02, 1.04] | <0.001 |
| CMS ownership type - for profit | 1.02 [1.02, 1.03] | <0.001 |
| CMS ownership type - government | 1.02 [1.01, 1.02] | <0.001 |
| Count of active prescription medication orders | 1.01 [1.01, 1.01] | <0.001 |
| American Indian or Alaska Native | 1.01 [1.00, 1.02] | 0.07 |
| Active ADRD diagnosis | 1.01 [1.01, 1.01] | <0.001 |
| Payer - managed care | 1.01 [1.01, 1.01] | <0.001 |
| Northeastern United States | 1.01 [1.00, 1.01] | <0.001 |
| Count of active over-the-counter medication orders | 1.00 [1.00, 1.00] | <0.001 |
| Resident current BMI (/5 units) | 1.00 [1.00, 1.00] | <0.001 |
| Average BIMS score | 1.00 [1.00, 1.00] | <0.001 |
| Facility Five-star rating (from CMS Provider Info) | 1.00 [1.00, 1.00] | 0.0011 |
| Native Hawaiian or Other Pacific Islander | 1.00 [0.97, 1.03] | 0.96 |
| Days in facility (/30 days) | 1.00 [1.00, 1.00] | <0.001 |
| Facility number of beds (from CMS Provider Info) (/25 beds) | 1.00 [1.00, 1.00] | <0.001 |
| SVI - Overall percentile (for facility county) | 1.00 [1.00, 1.00] | 0.0078 |
| Resident average ADL scores | 1.00 [1.00, 1.00] | 0.17 |
| Count of active diagnoses (unique ICD-10 codes) | 1.00 [1.00, 1.00] | <0.001 |
| Active depression diagnosis | 1.00 [0.99, 1.00] | 0.013 |
| Western United States | 0.99 [0.99, 1.00] | 0.0033 |
| Active ADRD medication order | 0.99 [0.99, 1.00] | <0.001 |
| Active hypertension diagnosis | 0.99 [0.99, 0.99] | <0.001 |
| Resident age | 0.97 [0.97, 0.97] | <0.001 |
| Midwest | 0.97 [0.97, 0.98] | <0.001 |
| Payer - Medicaid | 0.97 [0.97, 0.98] | <0.001 |
| Active diabetes diagnosis | 0.97 [0.96, 0.97] | <0.001 |
| Payer - private | 0.94 [0.94, 0.95] | <0.001 |
| Payer - other | 0.87 [0.86, 0.88] | <0.001 |
